# High Burden of COVID-19 among Unvaccinated Law Enforcement Officers and Firefighters

**DOI:** 10.1101/2021.11.24.21266396

**Authors:** Alberto J. Caban-Martinez, Manjusha Gaglani, Lauren E.W. Olsho, Lauren Grant, Natasha Schaefer-Solle, Paola Louzado-Feliciano, Harmony L. Tyner, Sarang K. Yoon, Allison L. Naleway, Michael Smith, Brian E. Sokol, Karen Lutrick, Ashley L. Fowlkes, Jennifer Meece, Roger Noriega, Marilyn Odean, Andrew L. Phillips, Holly C. Groom, Kempapura Murthy, Laura J. Edwards, Katherine D. Ellingson, Young M.Yoo, Alexandra Cruz, Karley Respet, Matthew S. Thiese, Jennifer L. Kuntz, Spencer Rose, Louise S. Hadden, Joe K. Gerald, Josephine Mak, Damena Gallimore-Wilson, Jessica Lundgren, Kurt T. Hegmann, Kayan Dunnigan, Meredith G. Wesley, Edward J. Bedrick, Julie Mayo Lamberte, John M. Jones, Angela Hunt, Matthew M. Bruner, Kimberly Groover, Preeta K. Kutty, Addison C. Testoff, Lindsay B. LeClair, Jini M. Etolue, Mark G. Thompson, Jefferey L. Burgess

## Abstract

Law Enforcement Officers (LEOs), firefighters, and other first responders are at increased risk of SARS-CoV-2 infection compared to healthcare personnel but have relatively low COVID-19 vaccine uptake. Resistance to COVID-19 vaccine mandates among first responders has the potential to disrupt essential public services and threaten public health and safety. Using data from the HEROES-RECOVER prospective cohorts, we report on the increased illness burden of COVID-19 among unvaccinated first responders. From January to September 2021, first responders contributed to weekly active surveillance for COVID-19-like illness (CLI). Self-collected respiratory specimens collected weekly, irrespective of symptoms, and at the onset CLI were tested by Reverse Transcription Polymerase Chain Reaction (RT-PCR) assay for SARS-CoV-2. Among 1415 first responders, 17% were LEOs, 68% firefighters, and 15% had other first responder occupations. Unvaccinated (41%) compared to fully vaccinated (59%) first responders were less likely to believe COVID-19 vaccines are very or extremely effective (17% versus 54%) or very or extremely safe (15% versus 54%). From January through September 2021, among unvaccinated LEOs, the incidence of COVID-19 was 11.9 per 1,000 person-weeks (95%CI=7.0-20.1) compared to only 0.6 (95%CI=0.2-2.5) among vaccinated LEOs. Incidence of COVID-19 was also higher among unvaccinated firefighters (9.0 per 1,000 person-weeks; 95%CI=6.4-12.7) compared to those vaccinated (1.8 per 1,000; 95%CI=1.1-2.8). Once they had laboratory-confirmed COVID-19, unvaccinated first responders were sick for a mean±SD of 14.7±21.7 days and missed a mean of 38.0±46.0 hours of work. These findings suggest that state and local governments with large numbers of unvaccinated first responders may face major disruptions in their workforce due to COVID-19 illness.

## LETTER TEXT

Law Enforcement Officers (LEOs), firefighters, and other first responders are at increased risk of SARS-CoV-2 infection compared to healthcare personnel,^1,2^ but have relatively low COVID-19 vaccine uptake. COVID-19 was the leading cause of line-of-duty deaths among LEOs in the United States from January through November 2021, accounting for 276 (65.7%) of 420 deaths.^3^ Resistance to COVID-19 vaccine mandates among first responders has the potential to disrupt essential public services and threaten public health and safety. Using data from the HEROES-RECOVER prospective cohorts, we report on the increased illness burden of COVID-19 among unvaccinated first responders.

Detailed methods of the prospective HEROES-RECOVER cohorts in 6 States are published previously.^4-6^ From January to September 2021, first responders contributed to weekly active surveillance for COVID-19-like illness (CLI). Self-collected respiratory specimens collected weekly, irrespective of symptoms, and at the onset CLI were tested by Reverse Transcription Polymerase Chain Reaction (RT-PCR) assay for SARS-CoV-2. Socio-demographic characteristics, attitudes toward COVID-19 vaccines, and information on the duration of illness and missed work associated with COVID-19 were collected via electronic surveys.

Among 1415 first responders, 17% were LEOs, 68% firefighters, and 15% had other first responder occupations (**Table**). Mean age (±SD) was 41.3±9.7 years. Most participants were male (79%), White race (95%), and non-Hispanic (71%). Unvaccinated (41%) compared to fully vaccinated (59%) first responders were less likely to believe COVID-19 vaccines are very or extremely effective (17% versus 54%) or very or extremely safe (15% versus 54%). Only one-third (35%) of 1163 first responders who completed an attitude survey said they trusted what the government says about the COVID-19 vaccines; this percentage among 363 unvaccinated first responders was even lower (12%).

**Table.** Socio-demographic characteristics of first responders, their attitudes toward COVID-19 vaccines, and incidence and characteristics of laboratory-confirmed COVID-19 illness during the January to September 2021 study period

| Socio-Demographic Characteristics | Total | Unvaccinated | Fully Vaccinated<sup>a</sup> | Standard Mean Difference<sup>b</sup> |
| --- | --- | --- | --- | --- |
| **Total Participants<sup>c,†</sup>** | **1415** | **586 (41)** | **829 (59)** |  |
| **Cohort location, n (%)** |  |  |  |  |
| Arizona | 692 (49) | 265 (45) | 427 (52) | 0.63 |
| Florida | 519 (37) | 291 (50) | 228 (28) |  |
| Oregon | 42 (3) | 4 (1) | 38 (5) |  |
| Minnesota | 35 (2) | 2 (0) | 33 (4) |  |
| Texas | 34 (2) | 12 (2) | 22 (3) |  |
| Utah | 93 (7) | 12 (2) | 81 (10) |  |
| **Demographic Characteristics** |  |  |  |  |
| Male Sex, n (%) | 1113 (79) | 462 (79) | 651 (79) | 0.01 |
| White Race, n (%) | 1338 (95) | 554 (95) | 784 (95) | 0.00 |
| Hispanic, n (%) | 416 (29) | 190 (32) | 226 (27) | 0.11 |
| Age, Mean ± SD | 41.3 ± 9.7 | 38.8 ± 9.7 | 43.1 ± 9.4 | 0.46 |
| **Occupation, n (%)** |  |  |  |  |
| Firefighters | 964 (68) | 419 (72) | 545 (66) | 0.15 |
| Law Enforcement Officers | 238 (17) | 81 (14) | 157 (19) |  |
| Other First Responders<sup>d</sup> | 213 (15) | 86 (15) | 127 (15) |  |
| **Attitudes toward COVID-19 Vaccine** |  |  |  |  |
| Total who completed survey, n (%)<sup>e</sup> | 1163 (88) | 363 (73) | 800 (97) |  |
| Believe vaccine is very or extremely effective | 491 (42) | 61 (17) | 430 (54) | 1.01 |
| Believe vaccine is very or extremely safe | 489 (42) | 54 (15) | 435 (54) | 1.10 |
| Trust what government says about the vaccine | 406 (35) | 45 (12) | 361 (45) | 0.91 |
| **Incidence and Characteristics of Laboratory-confirmed COVID-19 Illness** |  |  |  |  |
| **Laboratory-confirmed COVID-19 illnesses** |  |  |  |  |
| Total COVID-19 illnesses | 184 | 160 | 24 |  |
| Prior to vaccine availability<sup>†</sup> | 103 (56) | 103 (100) | NA |  |
| During vaccine availability<sup>†</sup> | 81 (44) | 57 (70) | 24 (30) |  |
| **Laboratory-confirmed COVID-19 Incidence per 1,000 person-weeks (95% CI)<sup>f</sup>** |  |  |  |  |
| Firefighters | 4.0 (3.0-5.3) | 9.0 (6.4-12.7) | 1.8 (1.1-2.8) |  |
| Law Enforcement Officers | 2.8 (1.3-5.8) | 11.9 (7.0-20.1) | 0.6 (0.2-2.5) |  |
| Other First Responders | 5.0 (3.0-8.3) | 8.7 (4.6-16.4) | 2.9 (1.3-6.3) |  |
| Duration of COVID-19 illnesses, Mean days ± SD<sup>g</sup> | 12.1 ± 21.7 | 14.7 ± 21.7 | 9.6 ± 14.5 | 0.28 |
| Missed work due to COVID-19, Mean hours ± SD<sup>g</sup> | 26.7 ± 40.0 | 38.0 ± 46.0 | 11.9 ± 22.7 | 0.72 |

A total of 184 COVID-19 illnesses with RT-PCR-confirmed SARS-CoV-2 infection (COVID-19) were identified among first responders before and after COVID-19 vaccines became available in mid-December 2020. From January through September 2021, among unvaccinated LEOs, the incidence of COVID-19 was 11.9 per 1,000 person-weeks (95%CI=7.0-20.1) compared to only 0.6 (95%CI=0.2-2.5) among vaccinated LEOs. Incidence of COVID-19 was also higher among unvaccinated firefighters (9.0 per 1,000 person-weeks; 95%CI=6.4-12.7) compared to those vaccinated (1.8 per 1,000; 95%CI=1.1-2.8). Once they had laboratory-confirmed COVID-19, unvaccinated first responders were sick for a mean±SD of 14.7±21.7 days and missed a mean of 38.0±46.0 hours of work. Duration of COVID-19 illness and missed work were lower among fully vaccinated first responders.

During the study period, LEOs and firefighters who remained unvaccinated were 20- and 5-times more likely, respectively to become sick with COVID-19 than their colleagues who were fully vaccinated against COVID-19. On average, first responders were sick with COVID-19 for over 2 weeks and missed close to 40 hours of work due to their illness. Although this is among the largest prospective studies of its kind,^4-6^ the relatively small number of COVID-19 illnesses among fully vaccinated participants limited the precision of estimates and precluded models of incidence that could adjust for potential confounders. Nonetheless, these findings suggest that state and local governments with large numbers of unvaccinated first responders may face major disruptions in their workforce due to COVID-19 illness. Workplace requirements for employees to be vaccinated against COVID-19 have been established for healthcare workers. Given that COVID-19 vaccines have proven highly effective in controlling COVID-19 and its variants to date, during this public health emergency, state and local governments should consider vaccine mandates for first responders with a particular focus on LEOs, and regular COVID-19 testing or alternative work assignments for unvaccinated personnel. The low trust in government among first responders in this cohort additionally suggests a need to develop alternate strategies relying on trusted non-governmental information sources to increase vaccination rates.

## Data Availability

Data will be available by CDC when objectives of the
research are complete.

